# Genetic contributions to epigenetic-defined endotypes of allergic phenotypes in children

**DOI:** 10.1101/2024.10.03.24314618

**Authors:** Emma E. Thompson, Xiaoyuan Zhong, Peter Carbonetto, Andréanne Morin, Jason Willwerscheid, Cynthia M. Visness, Leonard B. Bacharier, Meyer Kattan, George T. O’Connor, Katherine Rivera-Spoljaric, Robert A. Wood, Diane R. Gold, Gurjit K. Khurana Hershey, Christine C. Johnson, Rachel L. Miller, Christine M. Seroogy, Edward M. Zoratti, Peter J. Gergen, Albert M. Levin, Matthew C. Altman, Tina Hartert, Matthew Stephens, Daniel J. Jackson, James E. Gern, Christopher G. McKennan, Carole Ober

**Affiliations:** Department of Human Genetics, University of Chicago, Chicago IL; Department of Mathematics & Computer Science, Providence College, Providence, RI; Rho Inc., Federal Research Operations, Durham, NC; Department of Pediatric Allergy, Immunology and Pulmonary Medicine, Monroe Carell Jr Children’s Hospital at Vanderbilt University Medical Center, Nashville TN; Department of Pediatrics, Columbia University Medical Center, New York NY; Pulmonary Center, Boston University School of Medicine, Boston MA; Department of Pediatrics, Washington University School of Medicine, St. Louis, MO; Department of Pediatrics, Johns Hopkins University, Baltimore MD; Department of Environmental Health, Harvard T.H. Chan School of Public Health; Channing Division of Network Medicine, Brigham and Women’s Hospital, Harvard Medical School, Boston, MA; Division of Asthma Research, Cincinnati Children’s Hospital and Department of Pediatrics, University of Cincinnati College of Medicine, Cincinnati OH; Department of Public Health Sciences, Henry Ford Health, Detroit MI; Division of Clinical Immunology, Icahn School of Medicine at Mount Sinai, New York, NY; Department of Pediatrics, University of Wisconsin School of Medicine and Public Health, Madison WI; Division of Allergy and Clinical Immunology, Henry Ford Health, Detroit MI; National Institute of Allergy and Infectious Diseases, NIH, Bethesda, MD; Center for Bioinformatics, Henry Ford Health, Detroit, MI; Systems Immunology Division, Benaroya Research Institute Systems and Department of Medicine, University of Washington, Seattle WA; Department of Medicine, Vanderbilt University School of Medicine, Nashville TN; Department of Statistics, University of Pittsburgh, Pittsburgh, PA

## Abstract

Background

Asthma is the most common chronic respiratory disease in children, but little is known about genetic contributions to its underlying endotypes. To address this gap, we studied the methylome, transcriptome, and genome from children with extensive phenotyping from birth.

**Methods:** We performed DNA methylation (DNAm) studies using the Asthma&Allergy array and RNA-sequencing in nasal mucosal cells from 284 children (age 11 years) in the Urban Environment and Childhood Asthma (URECA) birth cohort with genotypes from whole-genome sequencing. Using empirical Bayes matrix factorization on all CpGs on the array, we derived 16 DNAm signatures and tested for associations between phenotypes and gene expression. We then replicated results in two additional cohorts and estimated the heritability of phenotype-associated signatures using single-nucleotide polymorphisms (SNPs) associated with an allergic disease, and with CpGs and genes associated with the signatures.

**Findings:** Three DNAm signatures were associated with at least one phenotype: allergic asthma, allergic rhinitis, allergic sensitization (atopy), total IgE, exhaled nitric oxide, or blood eosinophils. The genes correlated with each of the three signatures were enriched in networks reflecting inhibited immune response to microbes, impaired epithelial barrier integrity, and activated T2 immune pathways. We replicated the signature-phenotype associations in two additional birth cohorts. The estimated joint SNP heritabilities of the signatures were 0.17 (p=0.0027), 0.30 (p=9.3x10^-7^), and 0.16 (p=9.0x10^-7^), respectively.

**Interpretation:** We identified three significantly heritable DNAm signatures defining asthma and allergy endotypes across diverse populations. Our study demonstrated that epigenetic patterning in airway mucosal cells reflects perturbations in underlying biological processes related to the development of asthma and allergic diseases in childhood.

## Introduction

Asthma affects over 300 million people worldwide and is the most common chronic respiratory disease in children.^1^ Asthma diagnosis is based on clinical features. including wheezing, chest tightness, and reversible airflow obstruction. However, significant heterogeneity among children with these shared symptoms reflects varying contributions of genetic and non-genetic factors, each of which may differentially promote distinct pathogenic mechanisms, or endotypes, of asthma. For example, early life wheezing illnesses during respiratory infections,^2^ due to impaired responses to respiratory viruses,^3^ may precede asthma diagnoses in some children. Sensitization to allergens may also precede the diagnoses of asthma,^2^ reflecting activated type 2 (T2) immune pathways that may ultimately give rise to co-morbid allergic diseases.^4^ Early life wheezing illnesses and atopy are both associated with defective barrier function in airway epithelial cells.^5^ These three mechanistic pathways, or endotypes, contribute significantly to the development of asthma and other allergic diseases in childhood.

Genome-wide association studies (GWAS),^6^ single-cell^7,8^ and bulk^9^ RNA-sequencing studies of airway cells, and epigenome-wide association studies (EWAS) of DNA methylation (DNAm) in airway epithelial and blood cells^10^ have affirmed a central role for epithelial expressed genes in the etiology of childhood-onset asthma, but the relative contributions of genetic and non-genetic factors to specific endotypes of asthma is less clear. Moreover, previous studies have largely focused on individual single-nucleotide polymorphisms (SNPs), genes, or methylated CpG sites, whereas perturbations of developmental pathways likely result from coordinated effects of genetic, transcriptional, and epigenetic variation. Thus, systems-level approaches that integrate across omic domains are needed to better define the molecular mechanisms that underlie endotype-specific trajectories of asthma and allergic diseases.

In this study, we leveraged DNAm and gene expression measured in nasal mucosal cells (which are largely epithelial) and whole-genome sequencing in children enrolled in the Urban Environment and Childhood Asthma (URECA) birth cohort study.^9,11^ We measured DNAm using the Asthma&Allergy array,^12^ which assays methylation levels for CpGs in genomic regions associated with asthma or allergic diseases and that overlap with genome annotations for predicted enhancers, transcription factor binding sites, and other gene regulatory elements. We used unsupervised multi-omic approaches to identify heritable DNAm signatures associated with allergic phenotypes and correlated with key asthma-and allergy-related transcriptional networks, and replicated associations in two independent cohorts. Finally, we demonstrated that the DNAm signatures were significantly heritable. Overall, our study demonstrated that epigenetic patterning in airway mucosal cells reflects perturbations in underlying processes associated with the development of asthma and allergic diseases.

## Methods

### Study Subjects

Our primary analyses were performed in the URECA cohort (N=284), with replication studies in the Infant Susceptibility to Pulmonary Infections and Asthma Following RSV Exposure (INSPIRE) cohort^13^ (N=524) and the Children’s Respiratory Environment Workgroup (CREW) consortium cohorts^14^ (N=722). In all cohorts, we used a cross-sectional design. The subjects were all unrelated. Demographic and clinical characteristics of the subjects from each cohort are described in **Table 1**. Additional details are in Supplemental Methods.

**Table 1.** Characteristics of URECA, INSPIRE, and CREW samples. See Supplementary Methods for details on the seven component CREW cohorts.

| Characteristic | URECA | INSPIRE | CREW |
| --- | --- | --- | --- |
| Nasal sample type | Mucosal | Mucosal | Lavage |
| Sample size (N) | 284 | 524 | 722 |
| Mean age (range), years | 11.1 (10.9-12.4) | 6.3 (5.0-7.9) | 17.4 (11.1-32.4) |
| Female sex | 46.8% | 47.0% | 49.1% |
| *Parent-Reported Race and Ethnicity* |  |  |  |
| Hispanic | 20.0% | 9.5% | 12.9% |
| Non-Hispanic Black | 71.8% | 18.1% | 44.3% |
| Non-Hispanic White | <1% | 65.4% | 36.0% |
| Other | 7.9% | 7.6% | 5.8% |
| *Clinical Assessment Sample Sizes* |  |  |  |
| Asthma | 91 (32.0%) | 52 (9.9%) | 113 (15.7%) |
| Allergic asthma | 63 (22.1%) | 22 (4.2%) | 61 (8.4%) |
| Non-allergic asthma | 27 (9.5%) | 23 (4.3%) | 20 (2.7%) |
| Atopy ($\geq 1$ +SPT) | 177 (62.3%) | 186 (35.5%) | 283 (37.8%) |

### Clinical Phenotypes and Definitions

We focused our studies on 11 clinical phenotypes measured at age 10 in URECA children: allergic and non-allergic asthma, allergic rhinitis, allergic sensitization (atopy), total serum immunoglobulin E (IgE), fractional exhaled nitric oxide (FeNO), blood eosinophil count, measures of lung function (FEV_1_, FVC, and FVC_1_/FVC), and body mass index (BMI). Five of these measures were available in the INSPIRE and CREW cohorts: nonallergic asthma, allergic asthma, atopy, FeNO, and blood eosinophil counts. Diagnostic criteria, phenotype definitions, and additional details are provided in Supplemental Methods.

### DNA Methylation and Gene Expression Studies

DNA from nasal mucosal cells (largely epithelium) was collected from 284 URECA^11^ children at age 11 years and from 524 INSPIRE^13^ children at ages 5-8 years. DNA from nasal lavage cells were collected from 722 CREW^14^ subjects between the ages of 11 and 33 years, including 64 URECA children who were not included in the nasal mucosal cell studies. DNAm was assessed using the Asthma&Allergy DNAm array.^12^ See Supplementary Methods for additional details.

RNA was extracted from the same nasal mucosal cells as used for DNAm studies in URECA children, as described.^9,12^

### Genotype Studies

Genotypes for QTL mapping in URECA were derived from whole-genome sequencing of DNA from peripheral blood cells, as described.^15^ CREW subjects (including URECA and INSPIRE) were genotyped with the Illumina Genome Diversity Array (GDA, 8v1.0) at the Center for Inherited Disease Research (CIDR). Details on quality control steps, ancestry PC estimations, and genotype imputation are described in Supplemental Methods. Imputed genotypes and ancestry PCs were used for GWAS and heritability studies.

### Statistical Analyses

As a data reduction step, we performed unsupervised factor analysis of the CpGs on the Asthma&Allergy array in URECA. The number of latent factors present in the data was first estimated using FALCO (**F**actor **A**na**l**ysis in **Co**rrelated Data),^16^ including array as a random effect and the following variables as fixed effects: collection site, ancestry principal components (PCs) 1-3, percent squamous and percent ciliated epithelial cells, plate, and DNA concentration. This estimated 16 latent factors. Empirical Bayes matrix factorization was then performed by applying the software *flashier*^17,18^ specifying 16 latent factors (see Supplementary Methods for details). We refer to the 16 DNAm-defined factors as “DNAm signatures.”

Associations between each signature and each clinical phenotype were tested using a linear mixed-effects model in R (v4.4.0), including plate, DNA concentration, site, percent ciliated and percent squamous cells, ancestry PCs 1-3, and sex as fixed effects and methylation array as a random effect. In URECA, we considered signature-level significance as p<0.0045 (0.05/11 phenotypes) and study-wide significance as p<0.00028 (0.05/[11 phenotypes × 16 signatures]). For replication studies, we scored the 16 signatures in each subject using the CpG scores derived from URECA (see Supplementary Methods) and corrected for multiple testing as described above.

We also tested for correlations between each of the 16 DNAm signatures and the 15,620 genes expressed in nasal mucosal cells from the 252 URECA children with both DNAm and RNA-seq data using linear regression (see Supplementary Methods). Significance was assessed by a false discovery rate (FDR) ≤0.05. Upstream regulator analyses for genes significantly correlated with the DNAm signatures were performed using all genes detected as expressed in nasal epithelial cells as the background (iPathwayGuide; Advaita Corp. 2019).^19^

### eQTL and meQTL Studies

SNPs on autosomes with minor allele frequency (MAF) ≥0.05 from whole-genome sequences^15^ were used for quantitative trait locus (QTL) studies of nasal mucosal gene expression (eQTL; n=252) and DNAm (meQTL; n=283) from URECA subjects using QTLtools^20^ (see Supplementary Methods for additional details). SNPs associated with the expression of at least one gene or with methylation of at least one CpG at FDR≤0.05 are referred to as eSNPs or meSNPs, respectively.

### Estimating Heritability of the Phenotype-Associated DNAm Signatures

We first used the 5,539,938 imputed autosomal SNPs (MAF≥0.05; imputation r^2^ 0.8) available in 1,188 URECA, INSPIRE, and CREW subjects to perform a GWAS of each DNAm signature including cohort ID, sex, cell composition, plate, cohort, and ancestry PCs 1-3 as fixed effects and genetic kinship as a random effect. We then estimated the proportion of variation in each signature explained by genetic variation (heritability) using three sets of SNPs: (i) SNPs from an allergic rhinitis GWAS (http://www.nealelab.is/uk-biobank/) that were fine-mapped at posterior inclusion probability ≥0.01 (see Supplementary Methods), (ii) meSNPs for CpGs in at least one of the phenotype-associated signatures, and (iii) eSNPs for genes correlated with at least one of the phenotype-associated signatures. See Supplementary Methods for additional details.

## Results

### Defining and Characterizing Networks of CpGs and Genes

As a first data-reduction step, we applied factor analysis coupled with an empirical Bayes approach to learn a reduced representation of the 37,256 CpGs. This reduced representation consisted of 16 factors (DNAm signatures), each of which captured a pattern methylation levels across the CpG sites (S1-S16; Supplementary Figure 1). The first four DNAm signatures (S1-S4) included >18,000 CpGs each and were assumed to capture non-specific variation with global effects on methylation levels. The remaining DNAm signatures (S5-S16) included between 487 and 6,654 CpGs (**Table 2**).

**Table 2.** Description of the 16 signatures and associations with clinical outcomes in URECA.^11^. Unadjusted p-values are shown in each cell with the direction of effect shown in parentheses. Bolded red font are p-values at study-wide level of significance (p<0.00028); bolded black font are p-values at signature-level significance (p<0.0045). The gene networks associated with S5, S8, and S16 reflect impaired viral response, impaired barrier function, and activated T2 immune responses, respectively (see Figure 2 and Supplementary Figures 2-7). ns, p>0.05

| Signatures | # CpGs | # Correlated Genes | #AS DMCs<sup>a</sup> | Asthma and Allergic Diseases |  |  | Allergic Sensitization (Atopy) |  | Type 2 Inflammation |  | Lung Function Measures |  |  | Obesity |
| --- | --- | --- | --- | --- | --- | --- | --- | --- | --- | --- | --- | --- | --- | --- |
|  |  |  |  | Non-allergic Asthma Dx Age | Allergic Asthma Dx Age | Allergic Rhinitis | Atopy (Prop. Positive SPT) | Total Serum IgE | FeNO | Blood Eos. Count | FEV<sub>1</sub> | FVC | FEV<sub>1</sub>/FVC | BMI |
| S1 | 32,795 | 6,258 | 171 | ns | ns | ns | 0.017 | ns | ns | ns | ns | ns | ns | ns |
| S2 | 19,529 | 3,559 | 148** | ns | 0.047 (-) | 0.024 (-) | 0.0070 (-) | ns | 0.057 (-) | 0.0077 (-) | ns | ns | ns | 0.0077 (-) |
| S3 | 17,966 | 4,586 | 113* | ns | ns | ns | ns | ns | ns | ns | ns | ns | ns | ns |
| S4 | 20,601 | 0 | 104 | ns | ns | ns | ns | ns | ns | ns | ns | ns | ns | ns |
| **S5** | **6,654** | **4,300** | **110\*\*** | 0.0056 (+) | **9.52E-05** (-) | **0.0035** (-) | **1.58E-12** (-) | **1.14E-10** (-) | **2.38E-04** (-) | **1.47E-11** (-) | ns | ns | ns | ns |
| S6 | 4,428 | 109 | 60** | ns | ns | ns | ns | ns | ns | ns | ns | ns | ns | ns |
| S7 | 1,875 | 0 | 12 | ns | ns | 0.025 (-) | ns | ns | ns | ns | ns | ns | ns | ns |
| **S8** | **4,067** | **4,946** | **72\*\*** | ns | ns | 0.020 (-) | **7.14E-04** (-) | 0.037 (-) | 0.024 (-) | 0.014 (-) | ns | ns | ns | ns |
| S9 | 1,381 | 22 | 30** | ns | ns | ns | ns | ns | ns | ns | ns | ns | 0.035 (+) | ns |
| S10 | 2,470 | 0 | 6 | ns | ns | ns | ns | ns | ns | 0.0089 (-) | ns | ns | ns | ns |
| S11 | 487 | 1 | 0 | ns | ns | ns | ns | ns | ns | ns | ns | ns | ns | ns |
| S12 | 1,308 | 0 | 1 | ns | ns | ns | ns | ns | ns | ns | ns | ns | ns | ns |
| S13 | 556 | 1 | 0 | ns | ns | ns | ns | ns | ns | ns | ns | ns | ns | ns |
| S14 | 5,495 | 292 | 33 | ns | ns | ns | ns | ns | ns | ns | ns | ns | ns | ns |
| S15 | 532 | 0 | 22** | ns | ns | ns | ns | ns | ns | ns | ns | ns | ns | ns |
| **S16** | **3,602** | **3,714** | **65\*\*** | ns | ns | **0.0041** (+) | **0.0011** (+) | **6.14E-05** (+) | 0.013 (+) | **1.10E-07** (+) | ns | ns | ns | ns |

<sup>a</sup> Allergic sensitization (AS) differentially methylated CpGs (DMCs) as defined in (Morin et al. 2023<sup>12</sup>) (FDR<0.05). Asterisks denote significant enrichment for AS DMCs; \* p=0.0014; \*\* p<1 × 10<sup>-5</sup>. Enrichment p-values were determined by permutation (see Supplementary Methods).

In our previous study using the Asthma&Allergy array, we conducted an EWAS of allergic sensitization (atopy) in the same high-risk URECA children as in this study, focusing on individual CpGs.^12^ This phenotype is a measure of specific IgE response to allergens,^21^ a crucial step in the development of childhood allergic asthma and allergic diseases.^2^ In that study, we identified 193 differentially methylated CpGs (DMCs) at an FDR≤0.05; replication studies in the general population–based INSPIRE^13^ cohort demonstrated that these associations were robust to age, ancestry, ascertainment, and geography. Among DNAm signatures 5-16, the atopy DMCs in URECA were enriched among the CpGs defining six signatures (**Table 2**).

### Correlations between DNAm Signatures, Clinical Outcomes, and Gene Expression

To further characterize each DNAm signature, we tested for associations between the 16 signatures and 11 clinical measures, including asthma (n=2), an allergic disease (n=1), atopy (n=2), type 2 inflammation (n=2), lung function (n=3), and obesity (n=1). Three signatures (S5, S8 and S16), which included 145 of the 193 (75.1%) atopy DMCs, were associated with at least one phenotype at signature-level significance (**Table 2**). The directions of effect of the associations were consistent within each of the three signatures. No other signature was significantly associated with the 11 clinical outcomes. S5 was associated with non-allergic asthma at p=0.0056, which did not reach signature-level significance, and its direction of effect was opposite of S5 associations with the allergic and T2 phenotypes. Lung function measures and BMI were not associated with any of the signatures. Overall, 10,813 of the 37,256 (29%) CpGs on the array were assigned to the three signatures (S5, S8, or S16), which were associated with allergic diseases, atopy, and markers of T2 inflammation. Of these CpGs, 72% (7,761) were specific to one of the three signatures (**Figure 1A**). The CpGs assigned to each signature are reported in DataFile1.

**Figure 1.**
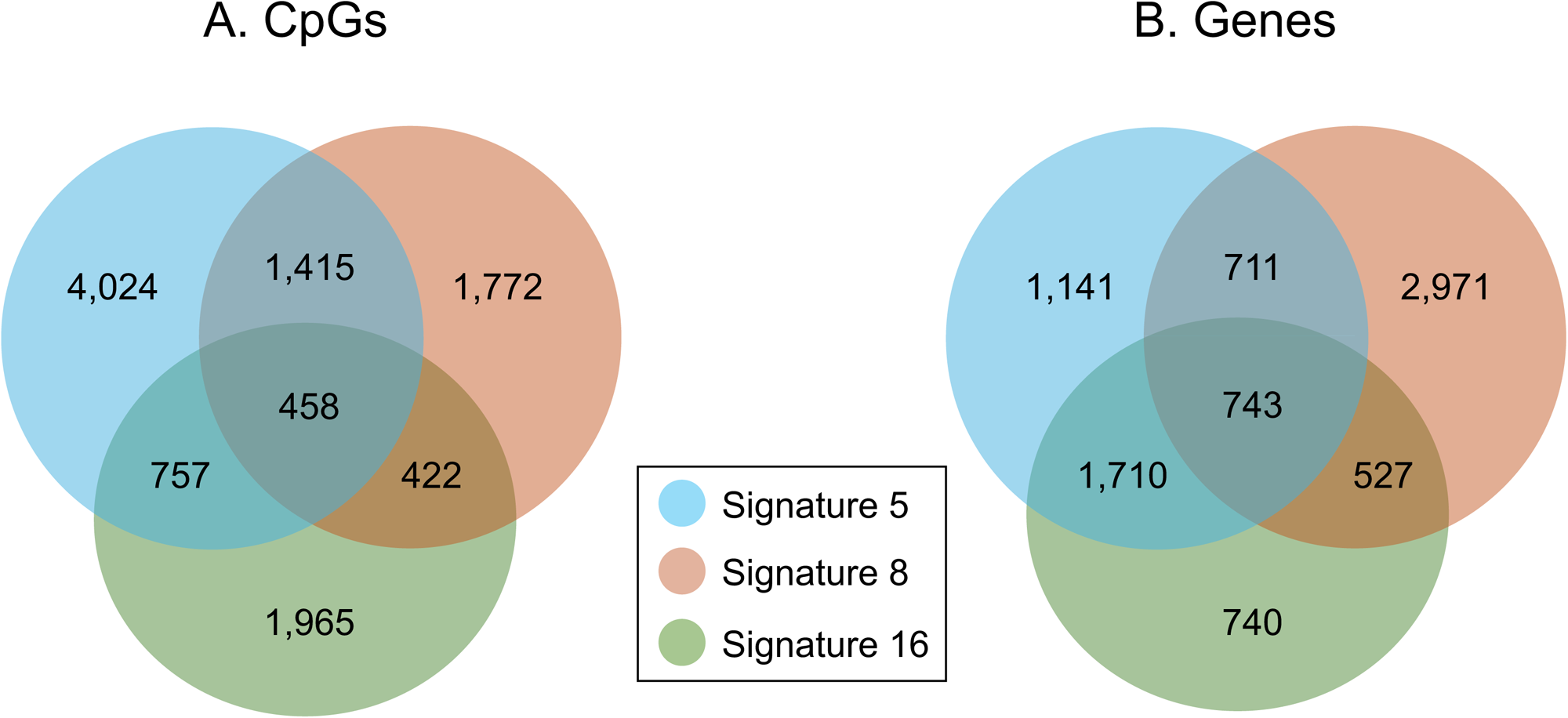
Overlap and number of CpGs and genes associated with DNAm signatures 5, 8, and 16. **A**) 7,761 out of 10,813 (71.8%) CpGs assigned to a signature (lfsr<0.05) are unique to only one signature; 4.2% are shared by all three. **B**) 4,852 out of 8,543 (56.8%) genes correlated (FDR<0.05) with at least one of the signatures are unique to only one signature; 8.7% are shared by all three.

To gain insight into the biology underlying the phenotype associations with DNAm signatures 5, 8, and 16, we tested for correlations between each signature and gene expression in the same airway mucosal cells that were used for DNAm studies, and then performed pathway analyses of each set of genes. Among the 15,620 expressed genes, 4,300, 4,946, and 3,714 genes were correlated S5, S8, and S16, respectively (FDR≤0.05) (**Table 2**). Of these genes, 56.8% (4,852) were specific to one of the three signatures (**Figure 1B**). The genes correlated with each signature are reported in DataFile2.

The two most significant upstream regulators of the genes correlated with S5 were *IFNG* (**Figure 2A** and Supplementary Figure 2A) and *IRF9* (Supplementary Figure 3). The expression of 19 of the 25 genes downstream of *IFNG* and all 16 of the genes downstream of *IRF9* were inhibited (i.e., more lowly expressed in URECA children with atopy). The two most significant upstream regulators of the genes correlated with S8 were *SEPTIN2* (**Figure 2B** and Supplementary Figure 2B) and *BBS1* (Supplementary Figure 4). The expression of all 47 genes downstream of *SEPTIN2* and all 8 genes downstream of *BBS1* were inhibited. The four most significant upstream regulators of the genes correlated with S16 were *IL13* (**Figure 2C** and Supplementary Figure 2C), *TSLP* (Supplementary Figure 5), *IL4* (Supplementary Figure 6), and *STAT3* (Supplementary Figure 7). The expression of 22 of 26, 8 of 9, and 32 of 42 genes downstream of *IL13*, *TSLP*, and *IL4*, respectively, were upregulated (i.e., more highly expressed in URECA children with atopy), whereas 36 of the 44 genes downstream of *STAT3* were inhibited.

**Figure 2.**
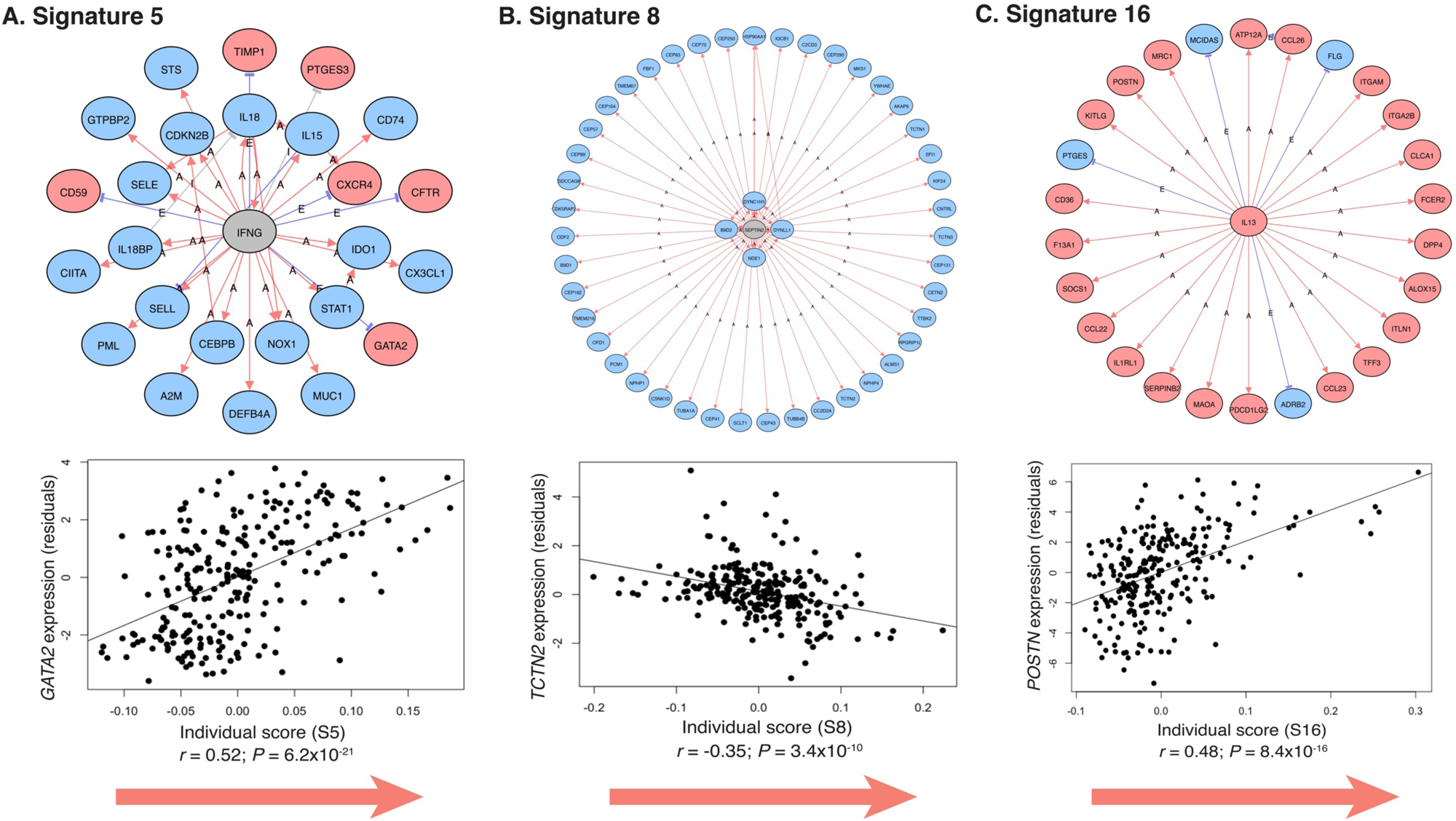
Significant upstream regulators of genes correlated with DNAm signatures 5, 8, and 16 (FDR<0.05). **A**) Interferon gamma (*IFNG*) is an upstream regulator of genes correlated with DNAm signature 5 (mean log_2_FC=-1.76; p=1.46 × 10^−4^). GATA binding protein 2 (*GATA2*) is downstream of *IFNG* and significantly activated in children with atopy. **B**) Septin-2 (*SEPTIN2*) is an upstream regulator of genes correlated with DNAm signature 8 (mean log_2_FC=-2.58; p=6.67 × 10^−16^). Tectonic family member 2 (*TCTN2*) is downstream of *SEPTIN2* and significantly inhibited in children with atopy. **C**) Interleukin 13 (*IL-13*) is an upstream regulator of gene correlated with DNAm signature 16 (mean log_2_FC=5.46; p=0.002). Periostin (*POSTN*) is downstream of *IL13* and significantly activated in children with atopy. Scatterplots show the correlation between expression of representative genes from each network (y-axis) and the individual scores for each DNAm signature (x-axis). The red arrow denotes the direction of the correlation with atopy in URECA subjects. The residuals shown in the scatterplots (y-axes) are after regressing out site, sequencing batch, ancestry PCs 1-3, and percent squamous and percent ciliated epithelial cells as covariates (Supplementary Methods). FDR-adjusted *P*-values are shown. Red symbols = activated genes; blue symbols = inhibited genes; gray symbols = not differentially expressed.

These results indicate that the three DNAm signatures associated with allergic and T2 phenotypes in the URECA children were correlated with independent sets of genes reflecting the inhibition of microbial response (S5) and impaired epithelial integrity (S8) pathways and the activation of T2 pathways (S16).

### Replication of Phenotype Associations in Other Cohorts

To assess the portability of the signatures defined by CpGs in the URECA children as predictors of allergic phenotypes more generally, we estimated the 16 signatures in 524 children in the INSPIRE cohort^13^ (**Table 1**) and used the same linear model as in URECA to test for associations between the 16 signatures and five phenotypes (study-wide significance p≤10^−4^; signature-wide significance p≤0.010). The three signatures were strongly associated with atopy in INSPIRE (p = 3.94 × 10^−18^ (S5), 2.76 × 10^−4^ (S8), and 5.38 × 10^−14^ (S16)) (Supplementary Table 1). S5 and S16 were also associated FeNO (8.3 × 10^-16^ and 4.6 × 10^-16^, respectively) and eosinophil count (3.4 × 10^-14^ and 6.9 × 10^-11^, respectively). S8 was also associated with eosinophil count (2.2 × 10^-4^) and nominally associated with allergic asthma (p=0.046). All associations showed the same direction of effect as that observed in URECA. S5 and S16 were associated with allergic asthma in the same direction of effect as in URECA but did not reach nominal significance in INSPIRE (p=0.10 and 0.091, respectively).

To further evaluate the generalizability to different airway cells and to older individuals, we performed DNA methylation studies using the Asthma&Allergy array in nasal lavage cells, comprised largely of airway immune cells, from 722 participants in CREW birth cohorts^11^ (**Table 1**). We tested for associations between the 16 signatures and five phenotypes (study-wide significance p≤10^−4^; signature-wide significance p≤0.010) (Supplementary Table 2). In these individuals, S5 was associated with allergic asthma (p=0.0082), atopy (p=8.22 × 10^−4^), FeNO (p=0.0027), and blood eosinophil counts (p=0.0031). S5 was nominally associated with non-allergic asthma (p=0.016) and, similar to URECA, with an opposite direction of effect compared to the allergic and T2 phenotypes. S16 was associated with FeNO (p=5.75 × 10^−5^) and eosinophil count (p=1.89 × 10^−6^) in the same direction as in URECA and INSPIRE. The associations of S16 with allergic asthma and atopy were not significant at nominal levels, but the directions of effect were consistent with the results in URECA and INSPIRE. The lack of association with S8 (reflecting impaired barrier function in URECA nasal mucosal cells) may be due to the small proportion of epithelial cells in nasal lavage samples.

These studies further indicate that DNAm signatures associated with inhibited immune response to microbes (S5) and activated T2 responses (S16) are shared with airway immune cells.

### GWAS and Heritability of DNAm Signatures 5, 8, and 16

We performed a GWAS of each DNAm signature in the 1,188 individuals in URECA, INSPIRE, and CREW with imputed genotypes. The GWAS showed no evidence of inflation (**Figure 3A**). Although no SNP associations were genome-wide significant after multiple testing correction in this small sample (p≤5 × 10^−8^), we observed enrichments of modest to small p-values among three groups of SNPs compared to null expectations: 537 likely-causal allergic rhinitis (AR) SNPs, 32,522 meSNPs for CpGs in S5, S8, or S16, and 384,030 eSNPs for genes correlated with S5, S8, or S16. AR SNPs and meSNPs were enriched for small GWAS p-values in all three signatures, while eSNPs were only enriched in the GWAS of S8 (**Figure 3A**).

**Figure 3.**
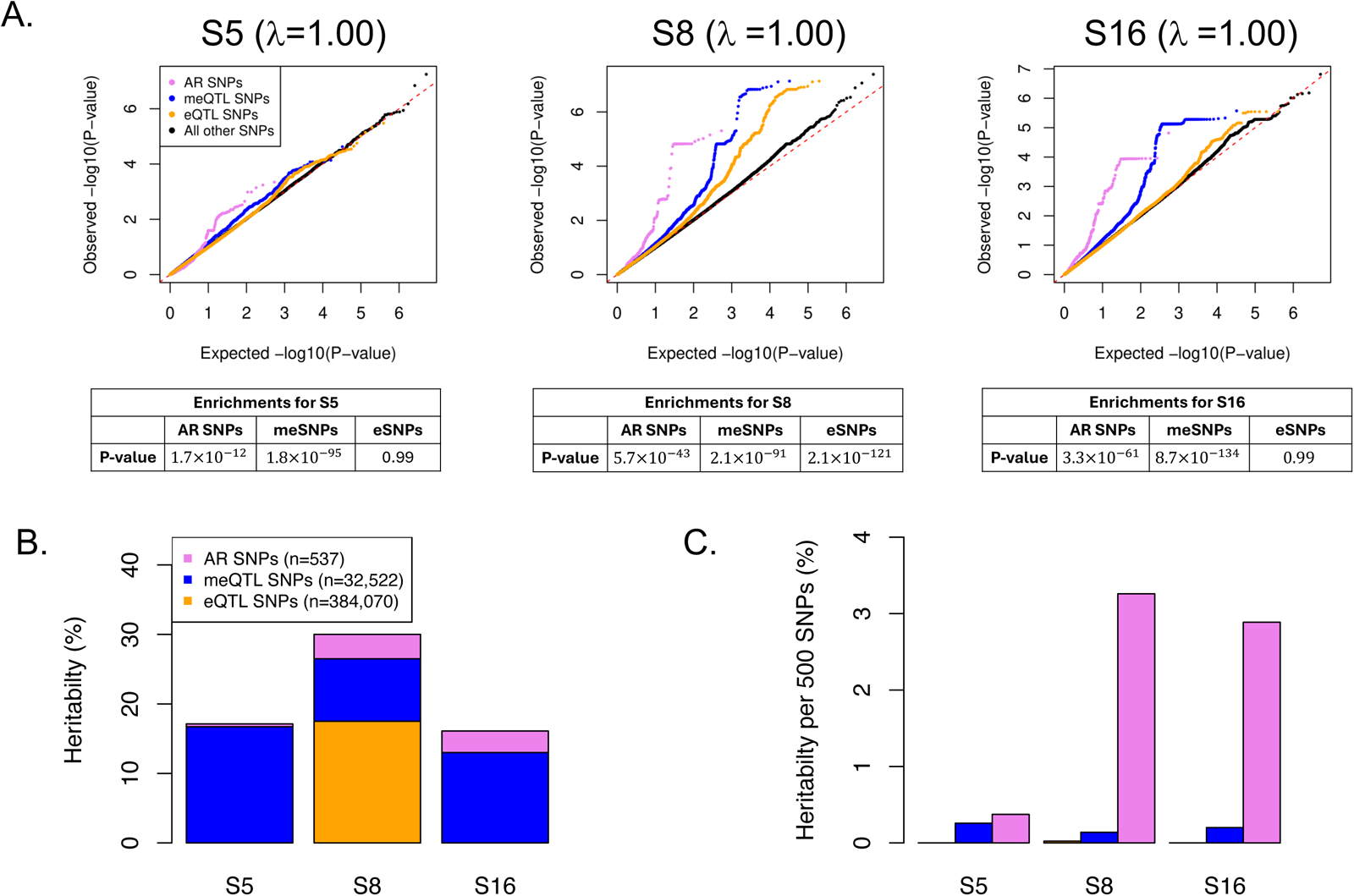
Heritability of the DNAm signatures. **A**) QQ plots of GWAS p-values (upper panel) and enrichment p-values (lower panel) for each signature. Genomic inflation factors, or lambdas (λ), were calculated based on all SNPs (black points). **B**) The contribution of each set of SNPs to the heritability of each signature. The total heritability of each signature was 0.17 (p=0.0026) for Signature 5, 0.30 (p=9.3x10^-7^) for Signature 8, and 0.16 (p=9.0x10^-7^) for Signature 16. **C**) The per-SNP contribution to the heritability of each signature.

Using these three sets of SNPs (n=417,089), the heritability of each DNAm signature was significant (**Figure 3B**), with meSNPs accounting for the largest portion of heritability in S5 and S16, and eSNPs accounting for the largest portion of heritability of S8. Because there were so many more meSNPs and eSNPs than AR SNPs in these analyses, we further estimated the per-SNP contributions to the heritability. AR SNPs had the largest per-SNP contribution to the heritability of all three signatures (**Figure 3C**).

## Discussion

Substantial epidemiologic evidence links prenatal and early life environments to the development of asthma and allergies in childhood, and GWAS affirm the important contribution of genetic variation to risk. Notably, DNAm measured in children and adults retains signatures of early life exposures^10,22,23^, suggesting that the methylome may provide an historical record of lifelong exposures. SNPs associated with DNAm levels (meSNPs) are highly enriched among those associated with asthma and allergic diseases in GWAS,^24–26^ possibly representing sites that mediate genotype-by-environment interactions. Therefore, DNAm provides an important framework for understanding the complex architecture underlying asthma and allergic diseases. Based on this premise, we performed an unsupervised multi-omic study to identify independent DNAm signatures in airway mucosal cells from 11-year-old children who were carefully phenotyped since birth, revealing insights into the epigenetic contributions to asthma and allergy endotypes that were not apparent in our previous EWAS of allergic sensitization in these same children.^12^

From among the 37,256 CpGs investigated, over one-third defined three signatures that were significantly associated with multiple different measures of allergic phenotypes and T2 inflammation and correlated with distinct transcriptional networks that reflected key endotypes of asthma. The replicated associations of signatures with allergic phenotypes in two independent cohorts reflect their robustness to age, ancestry, ascertainment, and geography. Two of the three signatures were common to both sources of upper airway cells (nasal mucosal cells, which are primarily epithelial, and nasal lavage cells, which are largely immune cells). None of the signatures were associated with pulmonary function measures or with BMI, indicating that in children the development of those phenotypes is independent of the nasal mucosal methylome and of the allergic and T2 features of asthma, consistent with clinical observations in URECA children showing that lung function and atopy in the first decade of life are largely independent risk factors for childhood asthma.^27^

All three signatures were associated with allergic asthma in at least one cohort and at least at nominal levels of significance. In contrast, only S5 was associated with non-allergic asthma in URECA (nasal mucosal cells) and CREW (nasal lavage cells) (p<0.05), both with opposite direction of effect compared to the allergic and T2 phenotypes in both cohorts. This finding supports the notion that allergic and non-allergic asthma have different underlying etiologies^28^ and further indicates that while allergic asthma may be associated with impaired immune responses to microbes, non-allergic asthma may be associated with a more activated response.

Our study further suggests that these differences are reflected in the nasal mucosal methylome of children and young adults with asthma.

While other studies have reported transcriptional signatures of asthma and allergic disease enotypes (e.g.^9,29–31^), this is the first demonstration of endotype-specific signatures define by DNAm. The most significant upstream transcriptional regulators of the genes correlated with each signature were specific to known asthma and allergy pathogenic programs, and the network genes correlated with S5 (inhibited response to microbes) and S16 (T2 immune activation) have been extensively studied in the context of asthma and allergy. In contrast, the genes in networks correlated with S8 (impaired barrier function) have not previously been implicated in these conditions. For example, S5 was correlated with networks of genes downstream of *IFNG* (interferon gamma) and *IRF9* (interferon regulatory factor 9), critical cytokines for activating innate and adaptive immune responses to viral and bacterial infections. The expression of nearly all those network genes were inhibited in the nasal mucosal cells of atopic URECA children (**Figure 2A**), consistent with an impaired response to airway microbes in children with allergic asthma, allergic rhinitis, or atopy.^3^

The most differentially expressed gene downstream of *IFNG* was *GATA2* (GATA2 binding protein 2), which was one of the few activated genes in the network. GATA2 is a transcription factor required for mast cell–mediated IgE responses to allergens.^32^ The increased expression of *GATA2* in a network of impaired interferon response reflects the opposing nature of the interferon and T2 cytokine responses,^33^ e.g., suppressing T2 with anti-IgE treatment (omalizumab) in children with moderate to severe asthma enhanced interferon responses to and reduced the severity of viral respiratory illnesses.^34^ All genes downstream of *IRF9* were inhibited; the most differentially expressed gene, *RSAD2* (radical S-adenosyl methionine domain-containing 2) plays a role in antiviral response and innate immune signaling^35^ (Supplementary Figure 3).

S16 was correlated with networks of genes downstream of signature cytokines for T2 inflammation in epithelial (thymic stromal lymhopoietin [*TSLP*]) and immune (interleukin 13 [*IL13*] and interleukin 4 [*IL4*]) cells, and for signal transducer and activator of transcript 3 (*STAT3*). Nearly all the genes in the T2 networks were activated in atopic URECA children. Among the most differentially expressed activated genes in each of these networks were *CCL2* (chemokine [CC motif] ligand 2), *POSTN* (periostin), and *SOCS1* (suppressor of cytokine signaling 1), respectively (**Figure 2C** and Supplementary Figures 5-7). It is notable that the three upstream regulators of activated genes correlated with S16, or their receptors, are targets of biologic therapies for T2 asthma and allergic diseases,^36^ highlighting the potential therapeutic relevance of the nasal mucosal methylome in identifying drug targets.

The genes downstream of *STAT3* were largely inhibited (Supplementary Figure 7). Among its many functions, STAT3 is an important determinant of whether naïve T cells differentiate into regulatory (Treg) or inflammatory (Th17) T cell lineages, which have overlapping developmental programs with Th1 and Th2 lineages.^37^ The inhibition of *STAT3* and its downstream genes in S16 may reflect the commitment toward T2 pathways and the downregulation of programs that favor Treg and Th17 cell differentiation. Consistent with this, macrophage inflammatory protein (MIP3A), encoded by the chemokine ligand 20 (*CCL20*) gene, is induced by the cytokine IL-17F in airway epithelial cells^38^ and was significantly inhibited in URECA children with atopy (Supplementary Figure 7).

The most significant upstream regulators of genes correlated with S8 were *SEPT2* (septin-2) and *BBS1* (Bardet-Biedel Syndrome 1) (**Figure 2B** and Supplementary Figure 4). SEPT2 is a component of the cytoskeleton^39^ and plays a significant role in bacterial invasion^40^ and barrier function.^41^ Fu et al. showed that SEPT2 has an IFN-gamma-independent effect on antiviral immunity and inhibits excessive inflammation in macrophages.^42^ Whether it plays a similar role in airway mucosal cells has not been explored, but the fact that it is a significant hub gene in a DNAm signature that is independent of the *IFNG*-mediated transcriptional networks in S5 suggests that *SEPT2* and its downstream genes may play a similar role in airway mucosal cells. Expression levels of all genes in this network were inhibited.

The most differentially expressed gene downstream of *SEPT2* was *TCTN2* (tectonic family member 2) (**Figure 2B**), a component of the cilia membrane that acts as a barrier. Absence of mouse *Tctn2* resulted in defects in ciliogenesis and ciliary membrane composition, and mutations in human *TCTN2* were associated with ciliary dysfunction diseases in humans.^43^ *BBS1* is one of seven BBS proteins that form the stable core of a complex required for ciliogenesis,^44^ and loss-of-function mutations in any one of these genes cause Bardet-Biedl syndrome, a prominent group of genetic ciliopathies. All genes downstream of *BBS1* were inhibited in atopic URECA children (Supplementary Figure 4). Interestingly, patients with Bardet-Biedl syndrome were reported to have an increased prevalence of practitioner-diagnosed asthma and rhinitis.^45^ Otherwise, there is a paucity of literature linking the genes in the S8 networks to asthma or allergic diseases. Our studies suggest that they may indeed play a critical role by both maintaining barrier integrity and preventing invasion and replication of microbes. These data also suggest that asthma and allergic diseases may represent another group of ciliopathies.^46^

We assumed that interindividual variation in the three DNAm signatures was due to both genetic variation and exposure histories. To tease apart these effects, we estimated their heritability. We further assumed that SNPs associated with AR in GWAS, meSNPs for the CpGs assigned to each signature, and eSNPs for the genes correlated with each signature would account for much of their heritability. Indeed, the heritability of each signature was significant. S8 (impaired epithelial integrity pathways) had the largest heritability (*h*^2^=0.30) and S5 (inhibited response to microbes) and S16 (activated T2 immune response) had smaller and similar heritabilities (*h*^2^=0.17 and 0.16, respectively). The contribution of AR SNPs to the heritability of S8 and S16 (**Figure 3B**) highlights the important roles of impaired epithelial integrity and T2 inflammation in this allergic disease,^47^ and the larger contributions of meSNP compared to eSNPs to the heritability of all three factors may reflect the importance of past exposures on remodeling of the mucosal methylome. These studies suggest that biases toward the development of inhibited immune response to microbes, impaired barrier integrity, and activation of T2 immune pathways are present at birth and mediate individual responses to the prenatal and postnatal environments during early development.

Although it is not feasible, or even possible, to measure all relevant environments during these development periods, the lasting effects of exposures on the epigenome can be measured. We assessed DNAm using the Asthma&Allergy array that targets CpG sites in regions of the genome previously associated with asthma or allergic diseases and with features consistent with gene regulatory functions.^12^ The relatively small number of CpGs on the array allowed for an unsupervised analysis by including all the CpGs in the factor analysis, which is not possible with other larger arrays. Our results suggested that discrete DNAm-associated mechanisms promote immune responses to microbes, barrier function integrity, and T2 immunity in the airway, consistent with results of an EWAS of nasal DNAm using the Illumina EPIC array.^48^

A limitation of our study was that it was conducted in samples collected at single timepoints. Therefore, we could not assess whether the DNAm signatures arose prior to the development of asthma, allergic, and T2 phenotypes or whether the DNAm signatures are merely reflecting the relative health or disease status of these children. The significant genetic contributions to the signatures suggest that at least 16% to 30% of the variation in DNAm signatures preceded the diagnosis of these conditions, but longitudinal studies in early life are required to determine the timing of development of the DNAm signatures relative to the onset of respiratory illnesses and atopic diseases and to the temporal development of each signature relative to each other. A second limitation was the relatively small sample size available for performing GWAS of the signatures, which motivated us to use a subset of all possible SNPs. Despite this, we were able to show that S5, S8, and S16 were significantly heritable and to partition the heritability between AR SNPs, meSNPs, and eSNPs. In fact, the relatively small number of SNPs likely led to underestimates of the true heritability of these signatures.

In summary, our study confirms a central role for the airway mucosal cell methylome in defining endotypes of asthma and allergic phenotypes and identified gene networks specific to those processes in diverse populations. We further quantified the relative contributions of genetic and non-genetic effects to the DNAm signatures and identified hub and downstream genes that may be considered as therapeutic targets for specific endotypes of asthma and allergic diseases.

## Supporting information

DataFile1

DataFile2

## Data Availability

DNA methylation and gene expression data from URECA are available via Gene Expression Omnibus (GEO) (GSE220874 and GSE145505, respectively). Genotype data for URECA are available at dbGaP (phs002921.v1.p1). DNA methylation data from INSPIRE are available at GEO (GSE267595). Submission of DNA methylation data to GEO and genotype data to dbGaP from CREW subjects are in progress and will be submitted prior to publication of this manuscript. Metadata for all cohorts will be submitted to GEO with the DNAm data.

## Contributors

EET, XZ, DJJ, JEG, CGM, and CO contributed to the conception or design of the study. CMV, LBB, MK, BRO, KR-S, RAW, DRG, GKKH, CCJ, RM, CS, EMZ, PG, MCA, TH, DJJ and JEG contributed patient samples and/or data. EET, EZ, PC, AM, JW, MS, and CGM oversaw or did the statistical analyses. AML performed genotype QC and imputation. EET, XZ, CGM, and CO wrote the initial draft of the manuscript. EET and CGM have accessed and verified the data. All authors reviewed or critically revised the manuscript for important intellectual content and gave final approval.

## Acknowledgements

The authors wish to thank the URECA, INSPIRE and CREW medical, nursing, and program staff; the children and families participating in the participating these studies; and Chris Reyes for providing editorial assistance with the preparation of the final manuscript. This work was supported by the US National Institutes of Health (NIH) (UG3 OD023282 to J.E.G., UM1 AI114271 to D.J.J., UM1 AI60040 to D.J., U19 AI095230 to C.O., R01 HG002585 to M.S. and R01 AG080590 to. C.G.M.). Members of the Children’s Respiratory and Environment Workgroup have also received NIH grant support for the: Childhood Asthma Study (R01AI024156, R03HL067427, R01AI051598), Cincinnati Childhood Allergy and Air Pollution Study (R01 ES11170, R01 ES019890), Columbia Center for Children’s Environmental Health Study (P01 ES09600, R01 ES008977, P30 ES09089, R01 ES013163, EPA R827027), Childhood Origins of Asthma study (P01 HL070831, U10 HL064305, R01 HL061879), Epidemiology of Home Allergens and Asthma Study (R01 AI035786), Urban Environment and Childhood Asthma study (NO1 AI25496, NO1 AI25482, HHS N272200900052C, HHS N272201000052I, NCRR/NIH RR00052, M01 RR00533, UL1 RR025771, M01 RR00071, UL1 RR024156, UL1 TR001079, UL1 RR024992, NCATS/NIH UL1TR000040), Infant Susceptibility to RSV Exposure Study (U19 AI095227, UG3UH30 OD023282, UL1 TR002243), Wayne County Health, Environment, Allergy, and Asthma Longitudinal Study (R01 AI050681, R56 AI050681, R01 AI061774, R21 AI059415, K01 AI070606, R21 AI069271, R01 HL113010, R21 ES022321, P01 AI089473, R21 AI080066, R01 AI110450, R01 HD082147). Members of the Children’s Respiratory and Environment Workgroup have also been supported by the Fund for Henry Ford Health System.

