## SupplementaryMaterials for "Genetic contributions to epigenetic-defined endotypes of allergic phenotypes in children"

### Table of Contents


| | | | Page |
|---|---|---|---|
| **Methods** | | | 3 |
| **Tables** | | | |
| | Supplementary Table 1. | Associations with clinical outcomes in INSPIRE | 8 |
| | Supplementary Table 2. | Associations with clinical outcomes in CREW | 9 |
| **Figures** | | | |
| | Supplementary Figure 1. | Percent variance explained by each signature | 10 |
| | Supplementary Figure 2. | Bar plots showing the magnitude and direction of effect for gene networks downstream of *IFNG* (signature 5), *SEPTIN2* (signature 8), and *IL13* (signature 16). | 11 |
| | Supplementary Figure 3. | *IRF9* is an upstream regulator of genes correlated with signature 5 | 12 |
| | Supplementary Figure 4. | *BBS1* is an upstream regulator of genes correlated with signature 8 | 13 |
| | Supplementary Figure 5. | *TSLP* is an upstream regulator of genes correlated with signature 16 | 14 |
| | Supplementary Figure 6. | *IL-4* is an upstream regulator of genes correlated with Signature 16 | 15 |
| | Supplementary Figure 7. | *STAT3* is an upstream regulator of genes correlated with signature 16 | 16 |
| **References** | | | 17 |

### Methods

#### *Cohort Descriptions*

The **URECA** (Urban Environment and Childhood Asthma) study enrolled pregnant women living in low-income neighborhoods in four U.S. cities (Boston, New York, Baltimore, St. Louis) between 2005 and 2007. The cohort consists of 560 children with at least one parent with an allergic disease or asthma.[1] Children with nasal mucosal DNAm were included in this study (n=284). The **INSPIRE** (Infant Susceptibility to Pulmonary Infections and Asthma Following RSV Exposure) study is a population-based observational birth cohort of 1,952 healthy infants in central Tennessee.[2] Infants were enrolled following delivery and followed longitudinally for the outcomes of asthma and allergic diseases. Children with available nasal mucosal DNAm results between ages 5-7 were included in this study (n=524). **CREW** (Children's Respiratory and Environmental Workgroup) is a consortium of 12 U.S. birth cohorts, including URECA and INSPIRE, with nearly 9,000 subjects who are diverse with respect to parent-reported race and ethnicity, geography, ascertainment, year of recruitment, and urban, suburban and rural lifestyles.[3] Seven of those cohorts (n=722) participated in the nasal lavage studies, including 64 URECA children who did not participate in the nasal mucosal study. The local institutional review board at each participating site approved the study protocol. Written informed consent or parent/guardian permission was obtained along with child assent as appropriate for participation in specific cohorts and the CREW protocol.

Characteristics of the CREW cohorts included in the nasal lavage study. Additional details and criteria for "high risk" in each cohort can be found in Gern et al. 2019[3]:

| CREW Cohort | Ascertainment | Age range at collection in years (median) | % female | N |
|---|---|---|---|---|
| **CAS:** Childhood Allergy Study | General | 29.4-32.4 (30.9) | 61.4 | 70 |
| **CCAAPS:** Cincinnati Childhood Allergy and Air Pollution Study | High risk | 15.9-18.1 (17.1) | 38.9 | 154 |
| **CCCEH:** Colombia Center for Children's Environmental Health | General | 16.2-21.3 (19.7) | 57.5 | 87 |
| **COAST:** Childhood Origins of ASThma | High risk | 19.1-20.7 (19.6) | 50.0 | 64 |
| **EHAAS:** The Epidemiology of Home Allergens and Asthma Study | High risk | 22.5-25.1 (18.1) | 56.2 | 32 |
| **URECA:** The Urban Environment and Childhood Asthma | High risk | 12.8-14.9 (13.2) | 57.8 | 64 |
| **WHEALS:** Wayne County Health, Environment, Allergy, and Asthma Longitudinal Study | General | 11.1-16.1 (13.9) | 50.6 | 251 |
| **Total Sample** | | **11.1-32.4 (15.1)** | **49.1** | **722** |

#### *Clinical Phenotypes*

All measurements in URECA were collected at age 10. Asthma was diagnosed based on symptoms, lung function, and physician diagnosis, as previously described.[4] Rhinitis was diagnosed based on symptoms (runny nose, stuffy nose, and sneezing).[5] Allergic asthma and allergic rhinitis was diagnosed based on the presence of asthma or rhinitis in addition to allergic sensitization. Allergic sensitization (atopy) was determined using skin prick testing to identify aeroallergen sensitization using a Multi-Test PC device (Lincoln Diagnostics, Decatur, IL)[3] and was defined as the proportion of positive SPTs to 14 airborne allergens. Body mass index (BMI) was calculated according to CDC guidelines, and z-scores were used in the analysis. Blood collected at annual clinic visits was processed at center-specific laboratories, and complete blood counts (CBC) with differentials were performed. Measurement of fractional exhaled nitric oxide (FeNO) was performed using the NIOX ((NIOX VERO®, Circassia Pharmaceuticals Inc., Morrisville, NC), employing a technique modified from Silkoff et al.[6] and following American Thoracic Society guidelines.[7] Lung function was was measured with spirometry following standard ATS criteria, as previously described.[8]

Allergic and non-allergic asthma, allergic sensitization, eosinophil count, and FeNO were defined in INSPIRE at ages 5-7 using the same criteria as in URECA.

In the CREW cohorts, asthma was defined as parent-reported doctor diagnosis at any time up until the assessment age, plus parental report of one or more wheezing episodes in the 12 months prior (Definition 2, in Visness et al.[9]). Subjects without a parent-reported doctor diagnosis of asthma were used as controls. Allergic sensitization was defined as the proportion of positive SPTs to nine perennial and seasonal allergens. Allergic and non-allergic asthma were defined as in URECA. Eosinophil counts were determined by a CBC with differentials, and FeNO was measured as described above. Measurements for both variables were taken at the visit nearest the age at sample collection; if measurements were missing at that timepoint, data from the previous visit were used. Due to the high correlation between allergic sensitization results at ages 5-7 and 10 years among subjects with both timepoints ($r$=0.62; p=$1.77\times10^{-6}$), data from ages 5-7 were used for correlations due to the smaller number of subjects with missing data, with the exception of WHEALS, for which allergic sensitization results were only available at age 10. See table below for the number of subjects per cohort with each clinical phenotype.

Number of subjects in each CREW cohort with phenotype data.

| CREW Cohort | N | Non-allergic asthma | Allergic Asthma | Allergic sensitization (SPT) | FeNO | Blood eosinophil count |
|---|---|---|---|---|---|---|
| CAS | 70 | 55 | 55 | 55 | 12 | 12 |
| CCAAPS | 154 | 138 | 138 | 139 | 141 | 143 |
| CCCEH | 87 | 0 | 0 | 0 | 79 | 82 |
| COAST | 64 | 63 | 63 | 63 | 39 | 38 |
| EHAAS | 32 | 20 | 20 | 20 | 27 | 29 |
| URECA | 64 | 61 | 61 | 62 | 21 | 64 |
| WHEALS | 251 | 132 | 132 | 141 | 170 | 166 |
| **Total** | **722** | **469** | **469** | **480** | **489** | **534** |

***Genotyping and Imputation in the CREW Cohorts***

CREW subjects were genotyped with the Illumina Genome Diversity Array (GDA, 8v1.0) at the Center for Inherited Disease Research (CIDR). Individuals were removed if they met any of the following quality control (QC) criteria: genotype vs. self-reported sex mismatches, unexpected duplicate samples, >10% missing genotypes, and autosomal heterozygosity more or less extreme five standard deviations (sd=0.8%) from the mean of 30.4%. SNPs meeting any of the following QC were excluded: >10% missing genotypes, exact Hardy-Weinberg p-value $<5\times10^{-8}$, or genotype concordance < 90%. A total of 1,785,122 directly genotyped SNPs remained following QC. Ancestry principal components were estimated from autosomal SNPs with MAF >0.005 and LD $r^2<0.10$, using the 1,000 genomes subjects as reference. Imputation was performed using the TopMed reference panel (r2 version 1.0.0) of >100,000 whole genome sequences (human genome build 38) and the TopMed Imputation Server (version 1.7.3). Specifically, Eagle (version 2.4) was used for genotype phasing and minimac4 (version 1.0.2) was used for genotype imputation. Genotype dosages for 5,539,938 SNPs with MAF > 0.05 and imputation r2 >0.80 in 1,188 CREW subjects with DNAm measures were included in the GWAS and heritability studies described below.

***DNAm Studies***

DNA from nasal mucosal cells (largely epithelium) was collected using nasal brushes from 284 URECA children at age 11 years and using flocked nasal swabs from 524 INSPIRE children at 5-6 years; samples were stored at –80°C. RNA and DNA were isolated from the swabs using the Qiagen AllPrep DNA/RNA kit. After quality control checks, 37,256 CpGs in URECA and 37,254 in INSPIRE (96.9% of CpGs on the array) were included in downstream analyses. See Morin et al.[10] for additional details regarding DNAm processing and QC in URECA and INSPIRE. DNA from nasal lavage cells were collected from 722 CREW[3] subjects between the ages of 11 and 33 years, including 64 URECA children who were not included in the nasal mucosal cell studies. The CREW DNAm data were processed using the same pipeline as in URECA and INSPIRE.[10] After quality control checks, 37,253 CpGs were included in downstream analyses.

Nasal lavage samples were collected from 722 subjects from seven CREW birth cohorts.[3] Samples were collected and processed from subjects ranging in age from 11 to 32 years, as previously described.[10]

***Empirical Bayes Factorization using DNAm data in URECA***

Empirical Bayes matrix factorization was performed by applying the software *flashier*[11,12] to the 37,256 × 264 matrix of DNAm residuals specifying 16 latent factors (see Manuscript). Flashier estimated a decomposition of the DNAm residuals matrix into two low-rank matrices (a matrix of CpG scores, with rows corresponding to CpG sites, and a matrix of individual scores, with rows corresponding to individuals). The priors on the elements of these two matrices were set to "adaptive shrinkage" priors[13]; that is, a mixture of point mass at zero plus and zero-centered Gaussians. These priors were adapted separately for each matrix, and for each factor k = 1, …, 16, using empirical Bayes methods. CpGs were assigned to factors by applying a local false sign rate (lfsr) of 0.05 to the elements of the CpG matrix.

***Estimating DNAm Signatures in Replication Cohorts: INSPIRE and CREW***

We projected samples in the replication cohorts onto the reduced representation learned from the discovery cohort by estimating the scores for replication samples using the CpG matrix estimated from the URECA discovery cohort. Briefly, *flashier*[12,14–15] assumes the $i$-th subject's DNAm profile $y_i$, which is a length $p$ = #CpGs vector whose entries are the M-values for each CpG in subject $i$, is

$$y_i = Lf_i + e_i,$$

where $L$ is the $p \times K$ "CpG matrix" for the $K$=16 factors, $f_i$ is a length-$K$ vector containing the scores for subject $i$, and $e_i$ is mean-0 Gaussian error. Given an estimate $\hat{L}$ for $L$ derived from URECA, we estimated $f_i$ in a replication sample using the ordinary least squares estimator $\hat{f}_i = \left(\hat{L}^\top \hat{L}\right)^{-1} \hat{L}^\top y_i$. We did not regress out nuisance covariates prior to estimating $f_i$ because we adjust for them in all downstream regressions involving $f_i$.

***Correlations with Gene Expression***

Correlations between expression of the 15,620 genes detected as expressed in the 252 URECA subjects with both DNAm data and RNA-seq data, and the 16 signatures were assessed using linear regression, including site, sequencing batch, ancestry PCs 1-3, and percent squamous and percent ciliated epithelial cells as covariates. Significance was assessed using a false discovery rate (FDR) of 5%.

***Enrichment of Allergic Sensitization DMCs in Signatures***

To assess enrichments of allergic sensitization (atopy) DMCs among the 16 signatures, we conducted 500,000 permutations for each signature in which we randomly sampled 193 CpGs (the total number of DMCs in Morin et al.[10]) from the total and recorded how many times the overlap was equal to or greater than the observed number of DMCs in that signature. The p-values were calculated as the number of times the permuted number exceed the observed, divided by 500,000.

***QTL Mapping***

Whole-genome sequencing data available in 283 URECA children[16] were analyzed using QTLtools,[17] excluding genes on the X chromosome. To determine the eQTL window size, a ±1 Mb window was used as the baseline, and subsequent iterations of QTLtools were run with decreasing window sizes. Our final window size of ±125 kb retained >90% of the number of FDR-adjusted significant SNP-gene pairs identified in the ±1 Mb window. For meQTL mapping, we used a window size of ±10 kb.

For eQTL studies, sex, ancestry PCs 1-3, site, percent squamous and percent ciliated cells, sequencing batch, and 21 expression PCs were included as covariates. For meQTL studies, sex, ancestry PCs 1-3, percent squamous and percent ciliated epithelial cells, and 14 methylation PCs were included as covariates. The number of expression PCs or methylation PCs in each analysis were estimated using FALCO[18], after regressing our technical and biological covariates.

We adjusted for expression and DNAm PCs in the eQTL and meQTL analyses to improve power, reduce biases, and ensure signatures are independent of eQTL and meQTL estimates. To see why the latter holds, since *flashier* and principal component analysis assume the same latent factor model for the data, signatures estimated by *flashier* are approximately equal to DNAm sample PCs after a change of basis (i.e., after rotating and scaling sample PCs). Therefore, adjusting for DNAm sample PCs in the QTL anlayses is equivalent to performing the analysis conditional on the signatures. As such, any statistical error in QTL estimates is approximately independent of signatures. We did not adjust QTL analyses for both PCs and *flashier* signatures to avoid overcorrecting and issues that arise with correlated columns in design matrices.

***Statistical Fine-Mapping of Allergic Rhinitis GWAS***

We performed Bayesian statistical fine mapping for an allergic rhinitis GWAS (http://www.nealelab.is/uk-biobank) of UK Biobank (UKB) European ancestry unrelated individuals (20,904 cases, 70,883 controls). First, we divided the genome into 1,703 approximately independent linkage disequilibrium (LD) blocks[19] computed on 1000 Genomes[20] European populations and selected LD blocks that contained at least one genome-wide significant (p < 5

$\times 10^{-8}$) SNP (significant blocks). We then used a modification of the "Sum of Single Effects" (SuSiE) model,[21] SuSiE-RSS,[22] to fine-map all significant blocks, allowing up to 5 independent causal signals per block. SuSiE-RSS takes GWAS z-scores and correlations of SNPs in a LD block as input and estimates the probability of each SNP being causal (i.e,, the posterior inclusion probability, or PIP). We computed the LD between SNPs on 10% of randomly sampled UKB British White individuals. Only SNPs with minor allele frequency greater than 1% were included in fine-mapping. Low-confidence variants suggested by the Neale Lab were removed from the fine-mapping analysis.

***Estimating Heritability of Signatures***

We used a random effects model to estimate the contribution of each group of SNPs (Group 1 = AR SNPs, Group 2 = meSNPs, and Group 3 = eSNPs) to the heritability of each signature. Briefly, let $n$ be the number of samples with genetic and DNAm data, $y$ be a length-$n$ vector whose entries are the signature value in each sample, $Z$ an $n \times r$ matrix of the $r$ nuisance covariates defined in Methods, and $X^{(j)}$ the $n \times p_j$ matrix of genotypes for the $j$-th group of SNPs. Missing genotypes in the URECA whole-genome sequencing data were replaced with the sample average. We standardized genotypes so that the genotype at each SNP had mean zero and variance one across samples by re-defining Group $j$'s genotype matrix to be $X^{(j)} \leftarrow P_Z^{\perp} X^{(j)} D^{(j)}$, where $P_Z^{\perp}$ is the orthogonal projection matrix that projects vectors onto the orthogonal complement of the image of $Z$ and $D^{(j)}$ is a $p_j \times p_j$ diagonal matrix whose $s$-th diagonal entry is the inverse of the sample standard deviation of the $s$-th genotype after regressing out $Z$. We then modeled $y$ as

$$y = Z\gamma + \sum_{j=1}^{3} X^{(j)}\beta^{(j)} + e, \qquad e \sim N(0, \sigma^2\{\rho K + (1-\rho)I\}), \qquad \rho \in [0,1],$$

where $K = 1/p_{\text{other}} X^{(\text{other})}\left\{X^{(\text{other})}\right\}^{\top}$ is the kinship matrix defined using all remaining genotypes $X^{(\text{other})}$, which was standardized in the same was as $X^{(j)}$. The kinship matrix accounts for the fact that some URECA subjects had both nasal brushing and lavage samples, as well as any cryptic relatedness between subjects. We took the standard approach to estimating heritability[23] by assuming $\beta^{(j)}$ was a random vector with independent entries distributed as $N(0, \phi_j^2/p_j)$. Under this model, the contribution to the heritability from the $j$-th group of SNPs is $\phi_j^2/(\sigma^2 + \sum_{j'=1}^{3} \phi_{j'}^2)$. Model parameters were estimated with restricted maximum likelihood (REML) and p-values for the null hypothesis that the heritability was zero, i.e., $H_0: \phi_1^2 = \phi_2^2 = \phi_3^2 = 0$, were computed assuming minus two times the log-likelihood ratio was distributed as $\frac{1}{8}\delta_0 + \frac{3}{8}\chi_1^2 + \frac{3}{8}\chi_2^2 + \frac{1}{8}\chi_3^2$ under the null hypothesis.

**Supplementary Table 1**. Associations with clinical outcomes in INSPIRE samples. P-values $<10^{-4}$ were study-wide significant and <0.010 were signature-wide significant. Directions of effect (+ or -) are shown in parentheses. Replicated associations with S5, S8, and S16 are shown in bolded red (study-wide significant) or black (signature-wide significant) font. ns, p>0.05

| | **Asthma** | | **Allergic Sensitization (Atopy)** | **Type 2 Inflammation** | |
|---|---|---|---|---|---|
| **Signature** | **Non-allergic Asthma** | **Allergic Asthma** | **Prop. Positive SPT** | **FeNo** | **Blood Eos. Count** |
| S1 | ns | ns | ns | ns | ns |
| S2 | ns | ns | ns | ns | 0.041 |
| S3 | ns | ns | 0.0015 | 0.022 | ns |
| S4 | ns | ns | ns | ns | ns |
| **S5** | ns | ns | **3.9E-18** (−) | **8.3E-16** (−) | **3.4E-14** (−) |
| S6 | ns | ns | 0.0086 (−) | ns | ns |
| S7 | ns | ns | ns | 0.016 | ns |
| **S8** | ns | 0.046 (−) | **2.8E-04** (−) | ns | **2.2E-4** (−) |
| S9 | ns | ns | ns | ns | ns |
| S10 | ns | ns | ns | ns | ns |
| S11 | ns | ns | **4.78E-4** (−) | ns | 0.032 |
| S12 | ns | ns | ns | ns | **7.6E-04** |
| S13 | ns | ns | **0.0028** (−) | ns | 0.041 |
| S14 | ns | ns | ns | ns | ns |
| S15 | ns | ns | ns | ns | ns |
| **S16** | ns | ns | **5.4E-14** (+) | **4.60E-16** (+) | **6.9E-11** (+) |

**Supplementary Table 2**. Associations with clinical outcomes in CREW nasal lavage samples. P-values <0.00091 were study-wide significant and <0.010 were signature-wide significant. Directions of effect (+ or -) are shown in parentheses. Replicated associations with S5, S8, and S16 are shown in bolded red font (study-wide significant) or bolded black font (signature-wide significant). ns, p>0.05

| | Asthma | | Allergic Sensitization (Atopy) | Type 2 Inflammation | |
|---|---|---|---|---|---|
| **Signature** | **Non-allergic Asthma Dx** | **Allergic Asthma Dx** | **Prop. Positive SPT** | **FeNo** | **Blood Eos. Count** |
| S1 | ns | ns | ns | ns | ns |
| S2 | ns | ns | ns | ns | ns |
| S3 | ns | ns | ns | ns | ns |
| S4 | ns | ns | ns | ns | ns |
| **S5** | 0.015 (+) | 0.011 (−) | **6.1E-04** (−) | **0.0027** (−) | **0.0037** (−) |
| S6 | ns | ns | ns | ns | ns |
| S7 | ns | ns | ns | ns | ns |
| **S8** | ns | ns | ns | ns | ns |
| S9 | ns | ns | ns | ns | ns |
| S10 | 0.032 (−) | ns | ns | ns | ns |
| S11 | ns | ns | ns | ns | ns |
| S12 | ns | ns | ns | ns | ns |
| S13 | ns | ns | ns | 0.046 (−) | ns |
| S14 | ns | ns | ns | ns | ns |
| S15 | ns | ns | ns | ns | ns |
| **S16** | ns | ns | ns | **3.9E-05** (+) | **5.2E-06** (+) |

**Supplementary Figure 1**. Proportion of variance explained (y axis) for each of the 16 signatures (x axis).

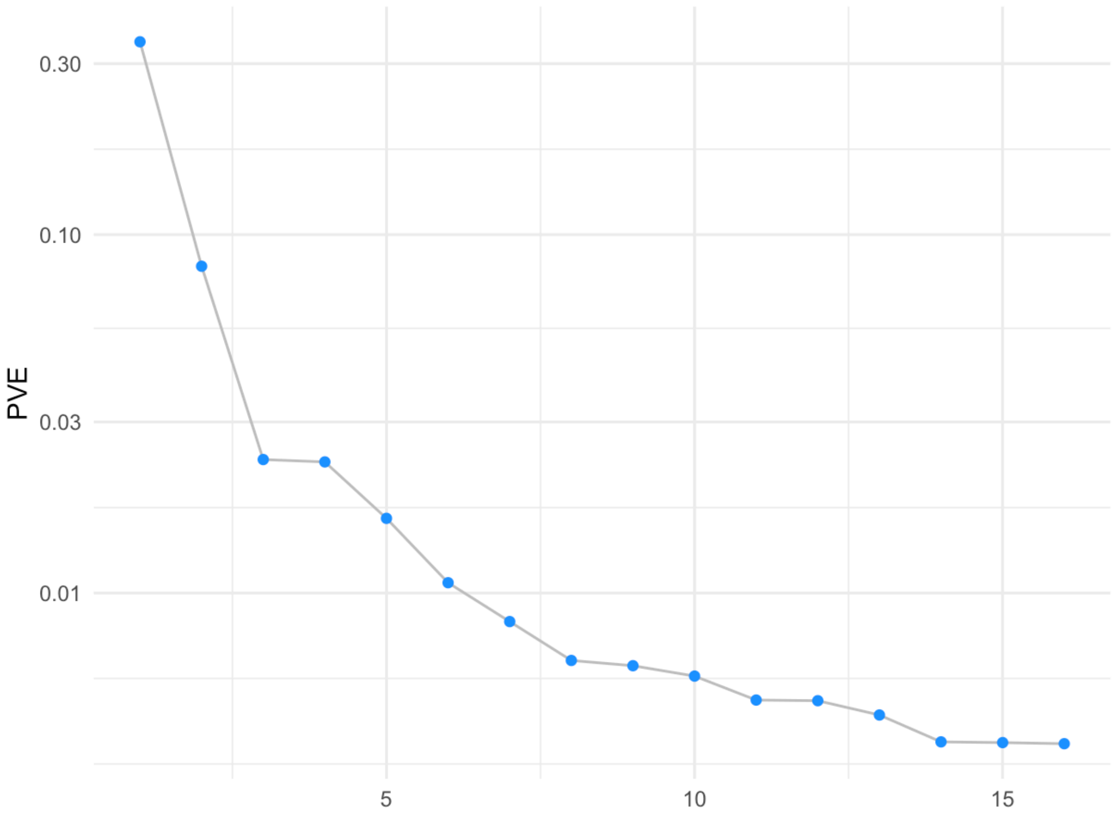

**Supplementary Figure 2.** Bar plots showing the magnitude and direction of effect for gene networks downstream of *IRF9* (Signature 5), *SEPTIN2* (Signature 8), and *IL13* (Signature 16). Boxplots on the left of each panel indicate the mean $\log_2$ fold change of genes in each network. Red symbols = activated genes; blue symbols = inhibited genes.

### A) Genes downstream of *IFNG* (Signature 5)

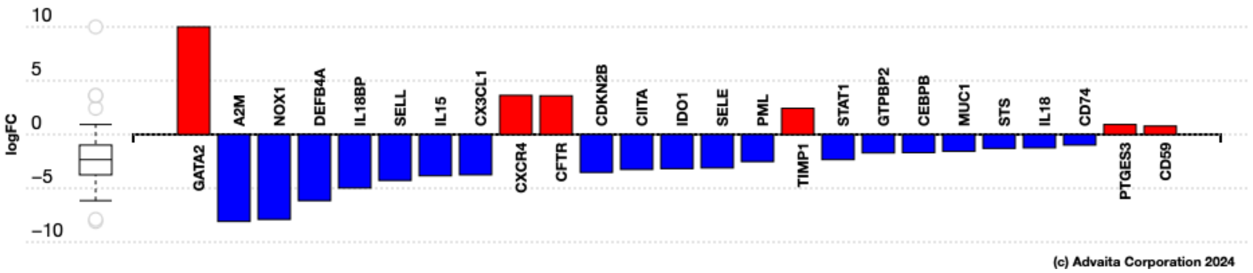


### B) Genes downstream of *SEPTIN2* (Signature 8)

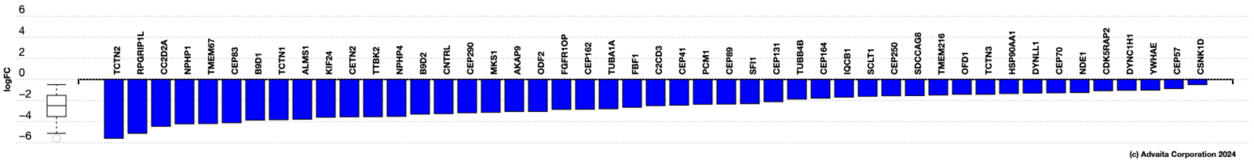


### C) Genes downstream of *IL13* (Signature 16)

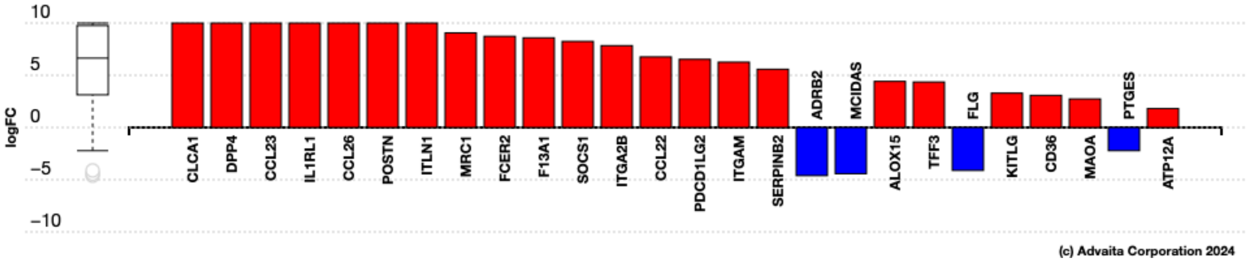

**Supplementary Figure 3**. *IRF9* (interferon regulatory factor 9) is an upstream regulator of genes correlated with signature 5 (impaired viral response) (mean $\log_2$ fold-change=-2.57; $P$=8.3x$10^{-5}$). A) Network of genes downstream of *IRF9*. B) Scatterplot showing the correlation between S5 (y-axis) and expression of *RSAD2* (radical S-adenosyl methionine domain containing 2). The red arrow denotes the direction of the correlation with allergic sensitization in URECA subjects. C) Bar plot showing the magnitude and direction of effect for genes in Panel A. Boxplot on the left of each panel indicates the mean $\log_2$ fold change of genes in each network. Blue symbols = inhibited genes

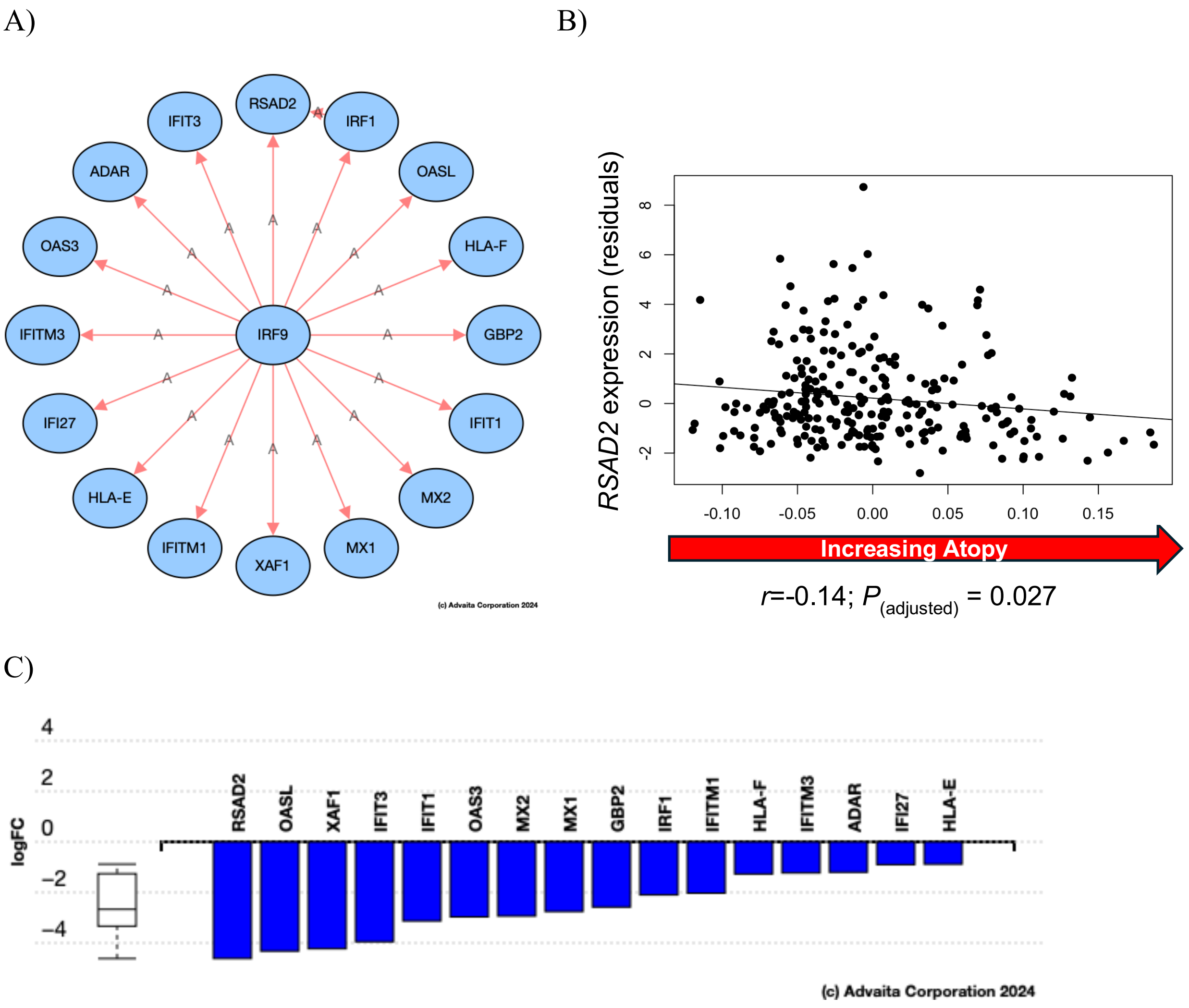

**Supplementary Figure 4**. *BBS1* (Bardet-Biedl syndrome 1) is an upstream regulator of genes correlated with signature 8 (impaired barrier function) (mean $log_2$ fold-change=-2.87; $P$=4.04x$10^{-6}$). A) Network of genes downstream of *BBS1.* B) The scatterplot shows the correlation between expression of *BBS2* (Bardet-Biedl syndrome 9) (y-axis), and signature 8 (x-axis). The red arrow denotes the direction of the correlation with allergic sensitization in URECA subjects.  C) Bar plot showing the magnitude and direction of effect for genes in Panel A downstream of *BBS1*. Boxplot on the left of each panel indicates the mean $log_2$ fold change of genes in each network. Blue symbols = inhibited genes; gray symbols = not differentially expressed.

A)

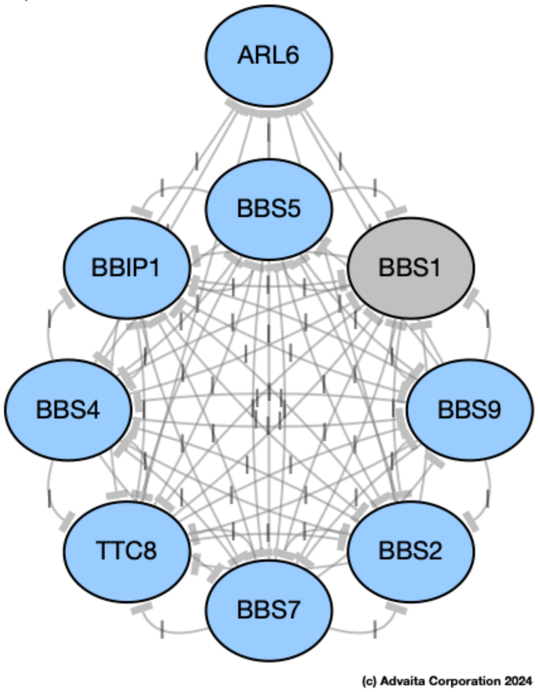


B)

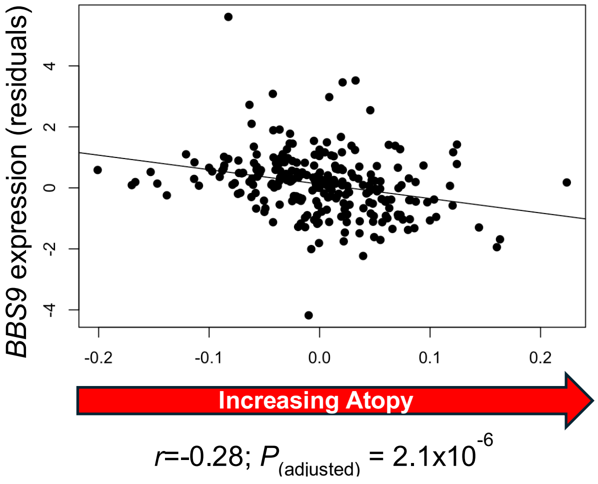


C)

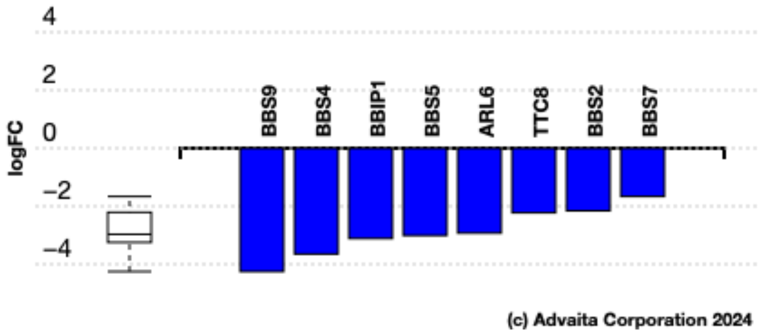

**Supplementary Figure 5**. *TSLP* (thymic stromal lymphopoietin) is an upstream regulator of genes correlated with signature 16 (T2 activation) (mean $\log_2$ fold-change=6.47; $P$=3.02x$10^{-4}$). A) Network of genes downstream of *TSLP*. B) The scatterplot shows the correlation between expression of *CCL2* (C-C motif chemokine ligand 2) (y-axis), and signature 16 (x-axis). The red arrow denotes the direction of the correlation with allergic sensitization in URECA subjects. C) Bar plot showing the magnitude and direction of effect for genes regulated by *TSLP*. Boxplot on the left of each panel indicates the mean $\log_2$ fold change of genes in each network. Red symbols = activated genes; blue symbols = inhibited genes; gray symbols = not differentially expressed.

A)

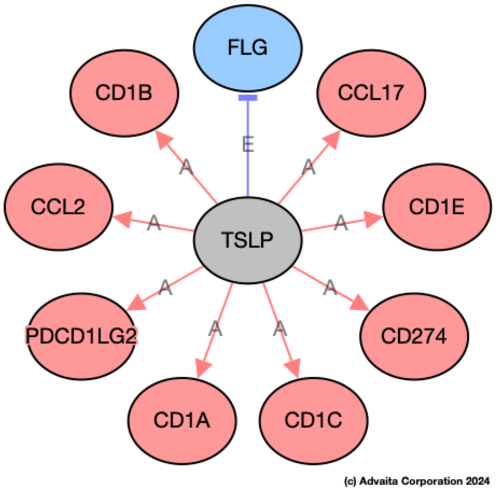


B)

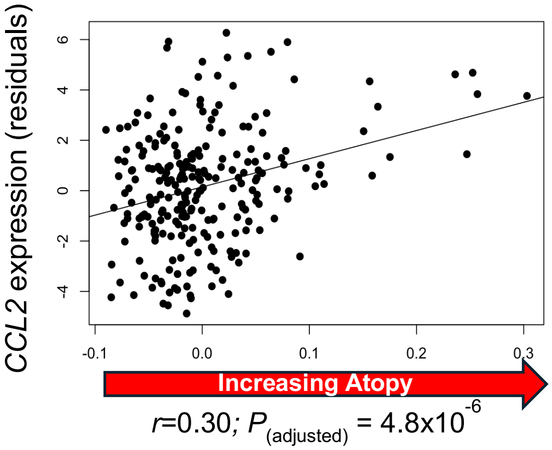


C)

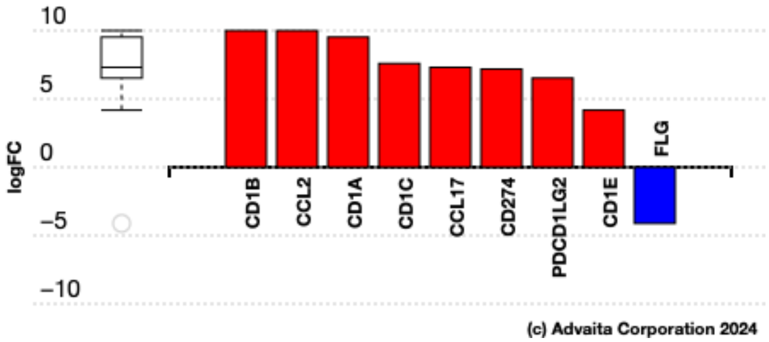

**Supplementary Figure 6**. *IL-4* (interleukin 4) is an upstream regulator of genes correlated with signature 16 (T2 activation) (mean $\log_2$ fold-change=4.22; *P*=0.001). A) Network of genes downstream of *IL4.* B) The scatterplot shows the correlation between expression of *SOCS1* (suppressor of cytokine signaling 1) (y-axis), and signature 16 (x-axis). The red arrow denotes the direction of the correlation with allergic sensitization in URECA subjects. C) Bar plot illustrating magnitude and direction of effect for genes regulated by *IL-4*. Boxplot on the left of each panel indicate the mean $\log_2$ fold change of genes in each network. Red symbols = activated genes; blue symbols = inhibited genes; white symbols = not expressed in nasal mucosal cells.

A)

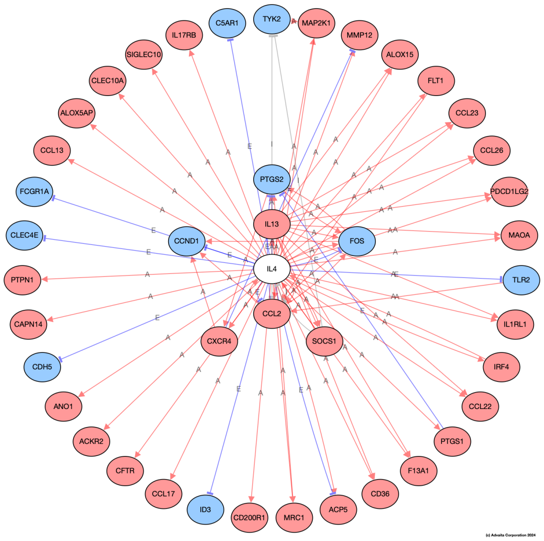


B)

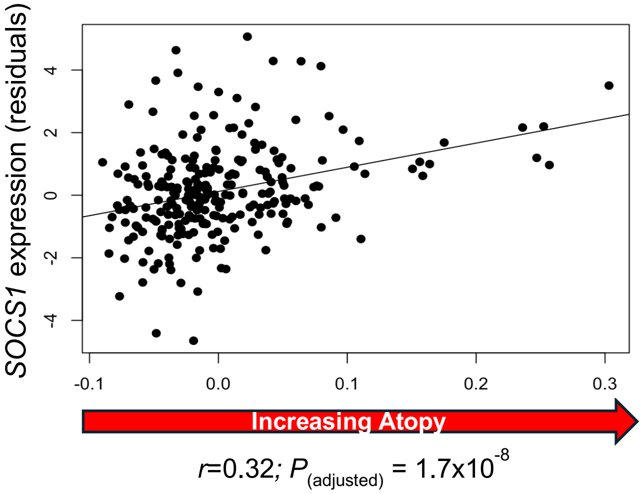


C)

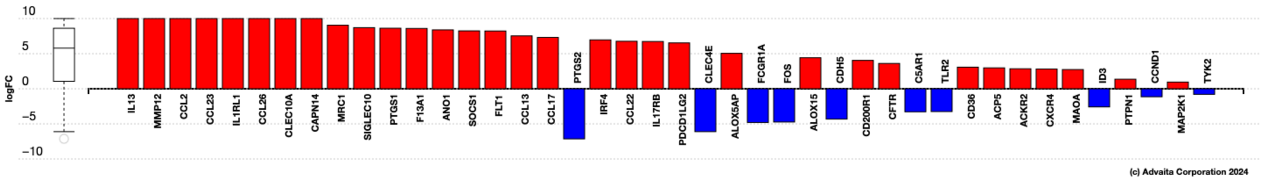

**Supplementary Figure 7**. *STAT3* (Signal transducer and activator of transcription 3) is an upstream regulator of genes correlated with signature 16 (T2 response) (mean $\log_2$ fold-change=-1.79; $P$=1.01x10$^{-4}$). A) Network of genes downstream of *STAT3.* B) The scatterplot shows the correlation between expression of *CCL20* (C-C motif chemokine ligand 20) (y-axis) and signature 16 (x-axis). The red arrow denotes the direction of the correlation with allergic sensitization in URECA subjects. C) Bar plot showing the magnitude and direction of effect for genes in Panel A regulated by *STAT3*. Boxplot on the left of each panel indicates the mean $\log_2$ fold change of genes in each network. Red symbols = activated genes; blue symbols = inhibited genes.

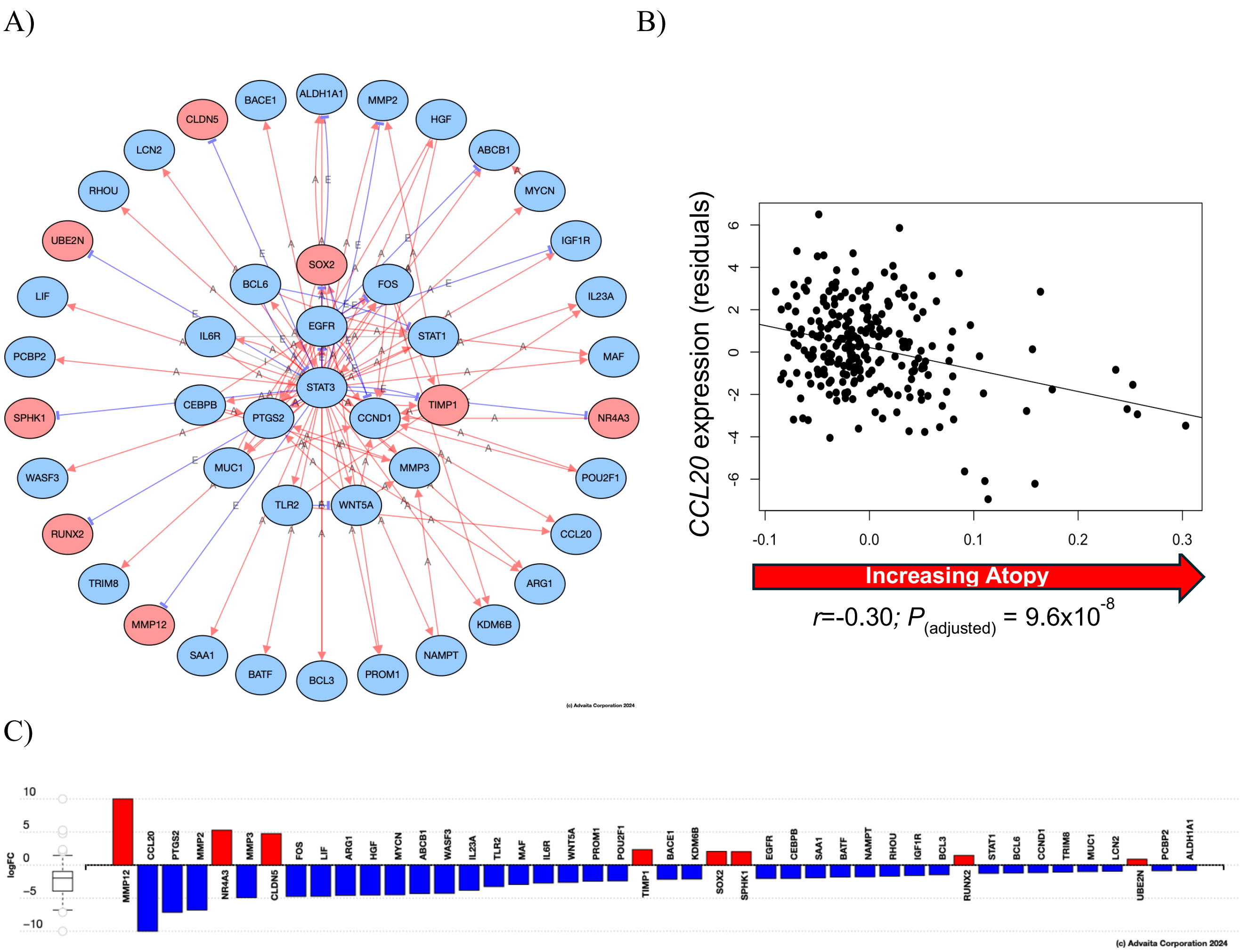
